# Temporal Trends and Associated Factors of Repeat Pregnancy in Ecuadorian Adolescents (2009–2022)

**DOI:** 10.1101/2025.05.27.25328420

**Authors:** Paola Toapanta-Pinta, Mercy Rosero-Quintana, Liseth Salazar-Congacha, Karla Benalcázar-Sanmartín, Santiago Vasco-Morales

## Abstract

**Objective:** To assess the temporal trend of repeat adolescent pregnancy in a tertiary-level hospital in Ecuador and identify associated maternal and perinatal factors.

**Materials and Methods:** A retrospective cross-sectional study using data from the Perinatal Information System of the Isidro Ayora Gynecological-Obstetric Hospital (2009-2022). Adolescent mothers aged 10 to 19 years were included. Temporal trends were analyzed, and sociodemographic, obstetric, and neonatal variables were compared between primigravida adolescents and those with repeat pregnancies. Binary logistic regression was applied to identify adjusted associations (p < 0.05).

**Results:** Of 6,695 adolescent births, 22.79% were repeat pregnancies. The rate declined between 2010 and 2014, which stabilized from 2016 to 2022 (20%-25%). Associated factors included age 15-19 years (aOR = 2.94; 95% CI: 1.96-4.41), lower education level (aOR = 1.55; 95% CI: 1.33-1.81), stable partner (aOR = 2.47; 95% CI: 2.16-2.81), planned pregnancy (aOR = 1.18; 95% CI: 1.02-1.35), exposure to violence (aOR = 1.55; 95% CI: 1.03-2.35), cesarean section (aOR = 1.22; 95% CI: 1.08-1.38), and preterm birth (aOR = 1.26; 95% CI: 1.06-1.50).

**Conclusion:** The stabilization of rates over the last decade suggests persistent structural barriers. Enhancing access to contraceptive services, comprehensive sexual education, and postpartum care is recommended to reduce repeat adolescent pregnancy.

## Introduction

Repeated adolescent pregnancy, defined as a second or more pregnancies before the age of 20, is a public health problem with significant medical, social, and economic consequences. During this crucial stage of development, early motherhood interrupts educational and work projects, perpetuating cycles of poverty and social vulnerability. ^1– 4^

Several factors contribute to repeat adolescent pregnancy, including adverse socioeconomic conditions, low educational attainment, early initiation of sexual activity, early unions, normalization of intimate partner violence, and limited contraceptive access.^5–8^In addition, some repeated pregnancies result from sexual abuse or coercion, which underscores the need for prevention strategies and comprehensive care.^3^ This type of pregnancy is associated with increased risks for the mother and newborn, including preterm birth, low birth weight, and obstetric complications. Despite their impact, repeat pregnancies tend to receive less attention than the first adolescent pregnancy, thus limiting the effectiveness of preventive strategies.^3, 9, 10, 11^

In Ecuador, the government has implemented policies to reduce early pregnancy in adolescents, including the Intersectoral Strategy for Family Planning and Prevention of Pregnancy in Adolescents (ENIPLA) 2011-2014,^12^ the National Plan for the Strengthening of the Family (2015) and the ^13^Intersectoral Policy for the Prevention of Pregnancy in Girls and Adolescents (Ecuador 2018-2025)^14^. However, the impact of these strategies on reducing repeat pregnancy remains uncertain.

Given the limited knowledge about this phenomenon in the Ecuadorian context, the present study aims to analyze the temporal trend of repeated pregnancy in adolescents in a national referral hospital and to identify the associated maternal and perinatal factors.

## Material and methods

A retrospective cross-sectional study was conducted based on records from the Perinatal Information System (SIP) of the Isidro Ayora Gynecological-Obstetric Hospital (HGOIA), in Quito, Ecuador, from January 2009 to December 2022. The HGOIA, a tertiary level hospital and national reference, has been providing comprehensive care to pregnant adolescents since 1988.

We included records of neonates born to adolescent mothers (10–19 years), excluding those with incomplete or inconsistent data. Maternal variables (age, education, ethnicity, marital status, gestational history), pregnancy factors (planning, contraceptive use, consumption of illicit substances, alcohol and tobacco, violence, anemia, premature rupture of membranes, hypertensive disorders, urinary tract infection, prenatal check-ups) and neonatal variables (type of delivery, being the product of multiple pregnancy, Apgar, gestational age, birth weight, congenital defects) and discharge status (alive, deceased, transferred).

Data were analyzed using R (version 4.2.0), employing the ‘Rcmdr’ and ‘EZR’ packages. The temporal trend of repeated pregnancy was evaluated, and a descriptive analysis was performed with absolute and relative frequencies. The comparison between first-time adolescents and adolescents with repeated pregnancy was made using the chi-square test. To identify the factors associated with repeated pregnancy, binary logistic regression was applied, calculating crude odds ratios (OR) and adjusted odds ratios (aOR) with their 95% confidence intervals (95% CI). The value of p < 0.05 was considered significant.

### Ethical considerations

The study was approved by the Human Research Ethics Committee of the Central University of Ecuador CEISH-UCE (code 009-DOC-FCM-2023). The data were anonymized, so no informed consent was required.

## Results

During the study period, 26,236 neonates were hospitalized at the HGOIA, of which 6,922 were children of adolescent mothers. After excluding 227 incomplete records or without information on the variable of interest, 6,695 cases were analyzed.

Of the total number of adolescent mothers, 22.79% (n=1,526) had at least one previous pregnancy. Among them, 3.64% (n=544) had a history of abortion and 16.79% (n=256) had undergone a previous cesarean section.

Repeat pregnancy rates declined from 2010 to 2014, then fluctuated between 15% and 25% from 2016 to 2022 (Figure 1).

**FIGURE 1.**
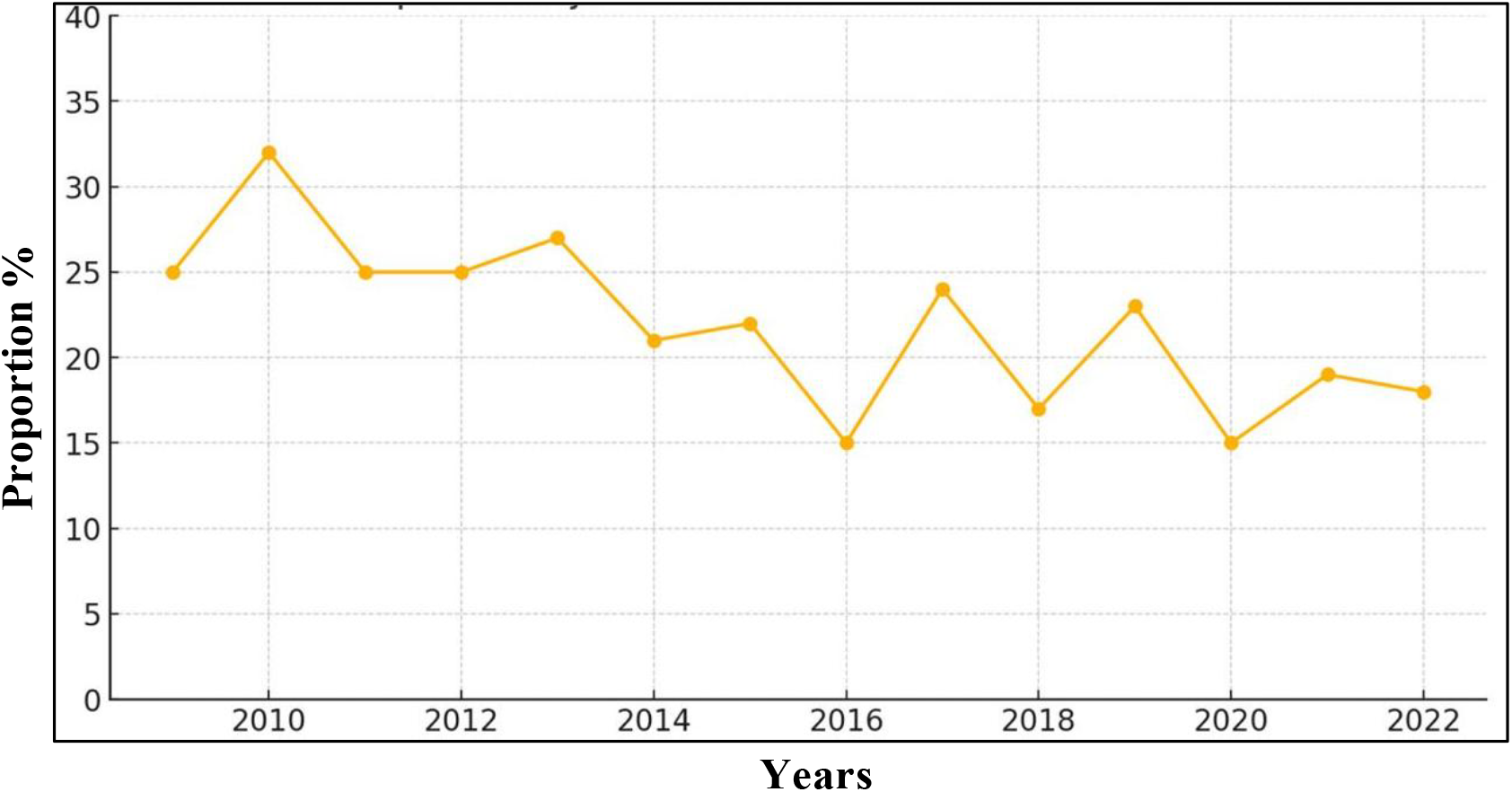
TREND IN THE PERCENTAGE OF REPEAT PREGNANCIES IN ADOLESCENTS BETWEEN 2009 AND 2022.

### Sociodemographic characteristics

Most adolescents were between 15 and 19 years old (95.13%), were mestizo (94.91%) and had reached secondary education (80.28%), 52.13% reported having a stable partner. Adolescents with repeated pregnancies were mainly older than 15 years (98.10% vs. 94.26%; p<0.001), with a higher proportion of primary education (22.15% vs. 14.76%; p<0.001) and more frequently had a stable partner (70.11% vs. 46.83%; p<0.001) (Table 1).

**Table I.**
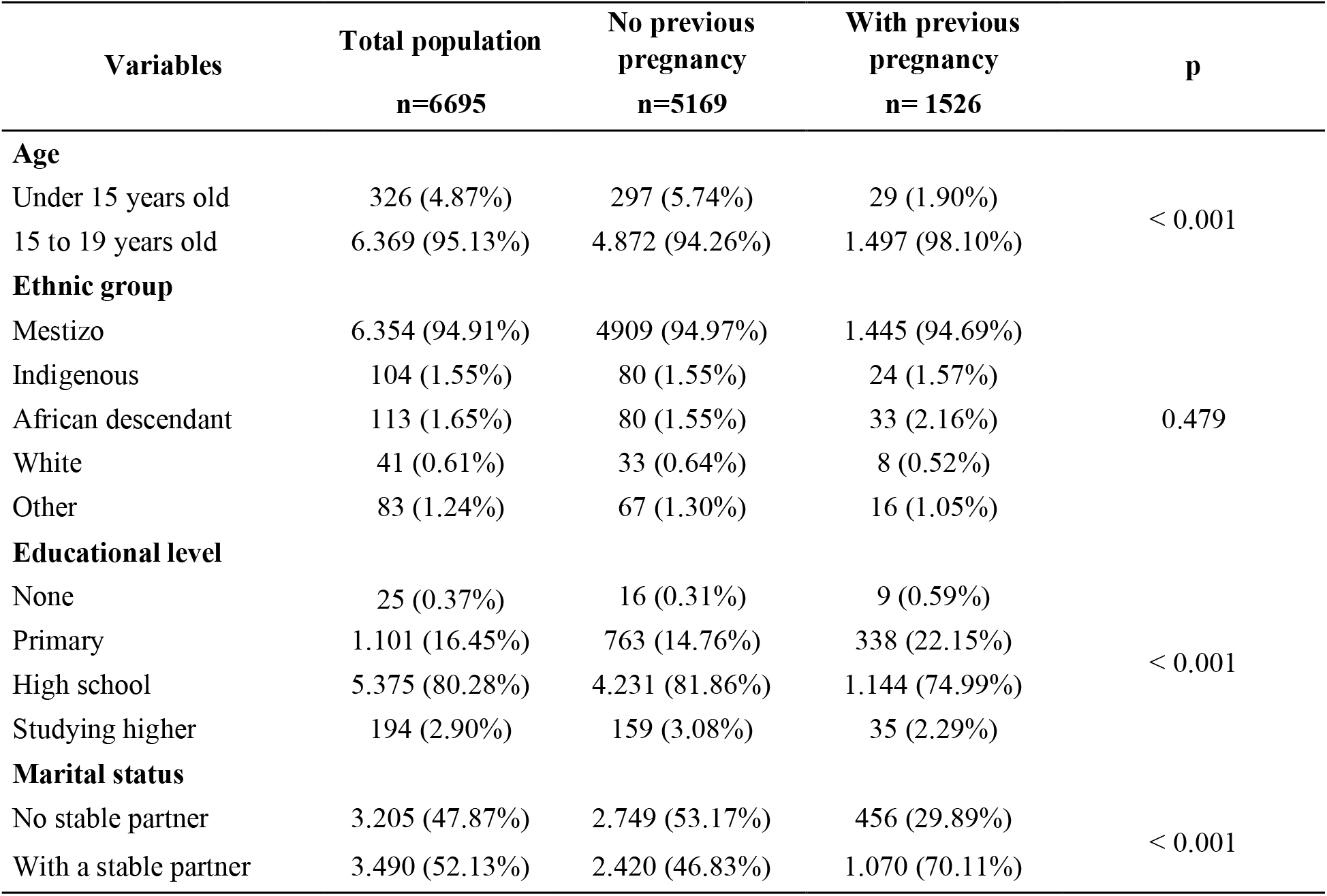
SOCIODEMOGRAPHIC CHARACTERISTICS AND COMPARISON OF GROUPS BETWEEN FIRST-TIME ADOLESCENTS AND ADOLESCENTS WITH REPEATED PREGNANCY. QUITO, ECUADOR, 2009-2022.

### Pregnancy-related factors

23.20% of pregnancies were planned and 9.75% were attributed to contraceptive failure. During pregnancy. 2.96% of adolescents consumed alcohol, 0.73% illicit substances and 6.54% were exposed to cigarettes. In addition, 1.94% reported having suffered violence (Table II).

**Table II.** PREGNANCY-RELATED VARIABLES AND GROUP COMPARISON BETWEEN FIRST-TIME ADOLESCENTS AND ADOLESCENTS WITH REPEATED PREGNANCY. QUITO. ECUADOR. 2009-2022.

| Variables | Total population<br>n=6695 | No previous<br>pregnancy<br>n=5169 | With previous<br>pregnancy<br>n= 1526 | p |
| --- | --- | --- | --- | --- |
| Planned pregnancy |  |  |  |  |
| Yes | 1.553 (23.20%) | 1.107 (21.42%) | 446 (29.23%) | < 0.001 |
| No | 5.142 (76.80%) | 4.062 (78.58%) | 1.080 (70.74%) |  |
| Contraceptive failure |  |  |  |  |
| Yes | 653 (9.75%) | 485 (9.39%) | 168 (11.01%) | 0.668 |
| No | 6.042 (90.24%) | 4.684 (90.61%) | 1.358 (89.00%) |  |
| Type of contraceptive * |  |  |  |  |
| Barrier | 340 (52.06%) | 254 (52.44%) | 86 (51.19%) | 0.774 |
| IUD | 4 (0.61%) | 2 (0.41%) | 2 (1.19%) |  |
| Emergency | 11 (1.68%) | 9 (1.85%) | 2 (1.19%) |  |
| Natural | 6 (0.91%) | 4 (0.82%) | 2 (1.19%) |  |
| Hormonal | 292 (44.77%) | 216 (44.53%) | 76 (45.23%) |  |
| Alcohol in pregnancy |  |  |  |  |
| Yes | 198 (2.96%) | 158 (3.06%) | 40 (2.62%) | 0.425 |
| No | 6.497 (97.04%) | 5.011 (96.94%) | 1.486 (97.37%) |  |
| Drug use |  |  |  |  |
| Yes | 49 (0.73%) | 37 (0.72%) | 12 (0.79%) | 0.909 |
| No | 6.646 (99.26%) | 5.132 (99.28%) | 1.514 (99.21%) |  |
| Tobacco use |  |  |  |  |
| Yes | 438 (6.54%) | 328 (6.35%) | 110 (7.21%) | 0.254 |
| No | 6.257 (93.45%) | 4.841 (93.65%) | 1.416 (92.77%) |  |
| Violence |  |  |  |  |
| Yes | 130 (1.94%) | 90 (1.74%) | 40 (2.62%) | 0.0371 |
| No | 6.565 (98.05%) | 5.079 (98.26%) | 1.486 (97.37%) |  |
| Anemia |  |  |  |  |
| Yes | 101 (1.51%) | 79 (1.53%) | 22 (1.44%) | 0.900 |
| No | 6.594 (98.49%) | 5.090 (98.47%) | 1.504 (98.56%) |  |
| RPM |  |  |  |  |
| Yes | 877 (13.09%) | 679 (13.14%) | 198 (12.98%) | 0.904 |
| No | 5.818 (86.90%) | 4.490 (86.82%) | 1.328 (87.02%) |  |
| Gestational hypertensive disorder |  |  |  |  |
| Yes | 372 (5.56%) | 299 (5.78%) | 73 (4.78%) | 0.151 |
| No | 6.323 (94.44%) | 4.870 (94.28%) | 1.453 (95.23%) |  |
| Urinary tract infection |  |  |  |  |
| Yes | 3.708 (55.38%) | 2.871 (55.56%) | 837 (54.88%) | 0.653 |
| No | 2.987 (44.61%) | 2.298 (44.56%) | 689 (45.14%) |  |
| Number of antenatal check-ups |  |  |  |  |
| None | 170 (2.54%) | 130 (2.52%) | 40 (2.62%) | 0.142 |
| 1 a 4 | 1.903 (28.40%) | 1.434 (27.72%) | 469 (30.74%) |  |
| 5 a 7 | 2.915 (43.54%) | 2.276 (44.08%) | 639 (41.86%) |  |
| 8 or more | 1.707 (25.50%) | 1.329 (25.70%) | 378 (24.78%) |  |

IUD (Intrauterine Device) . \* n:653

Adolescents with repeated pregnancy planned their pregnancy more frequently (29.32% vs. 21.42%; p<0.001). No significant differences were observed in alcohol, tobacco, or drug use. nor in maternal complications or number of prenatal check-ups between the two groups (Table II).

### Neonatal Outcomes

In the total population. 41.58% of births were by cesarean section. and 1.28% of newborns were the result of multiple pregnancies. At the first minute, 19.69% of newborns had an Apgar score <7, and 3.68% remained below 7 at the fifth minute, 35.34% of the newborns were preterm, 47% had low birth weight, and 33.56% of the newborns had low birth weight for their gestational age. There were 12.14% congenital defects and a neonatal mortality of 5.27%.

Compared to primiparous adolescents, those with repeat pregnancies had a higher frequency of cesarean deliveries (46.46% vs. 40.14%; p<0.001), preterm births (39.38% vs. 34.15%; p<0.001) and low birth weight (50.13% vs. 46.08%; p<0.001). There were no significant differences in Apgar scores, weight-for-gestational age, birth defects, or neonatal mortality between groups. (Table III).

**Table III.** CHARACTERISTICS OF CHILDBIRTH. NEONATAL OUTCOMES AND COMPARISON OF GROUPS BETWEEN FIRST-TIME ADOLESCENTS AND ADOLESCENTS WITH REPEATED PREGNANCY. QUITO. ECUADOR. 2009-2022.

| Variables | Total population<br>n=6695 | No previous<br>pregnancy<br>n=5169 | With previous<br>pregnancy<br>n= 1526 | p |
| --- | --- | --- | --- | --- |
| Type of delivery |  |  |  |  |
| Caesarean section | 2.784 (41.58%) | 2075 (40.14%) | 709 (46.46%) | < 0.001 |
| Vaginal | 3.911 (58.41%) | 3094 (59.85%) | 817 (53.54%) |  |
| Multiple Pregnancy Product |  |  |  |  |
| Yes | 86 (1.28%) | 65 (1.25%) | 21 (1.37%) | 0.538 |
| No | 6.609 (98.71%) | 5104 (98.74%) | 1505 (98.62%) |  |
| Apgar to the minute |  |  |  |  |
| < 7 | 1.317 (19.687%) | 1027 (19.86%) | 290 (19.00%) | 0.477 |
| ≥ 7 | 5.318 (80.32%) | 4142 (80.13%) | 1236 (81.00%) |  |
| Apgar at the fifth minute |  |  |  |  |
| < 7 | 247 (3.68%) | 189 (3.66%) | 58 (3.80%) | 0.852 |
| ≥ 7 | 6.448 (96.31%) | 4980 (96.34%) | 1468 (96.20%) |  |
| Gestational age |  |  |  |  |
| Preterm | 2.366 (35.34%) | 1765 (34.15%) | 601 (39.38%) | < 0.001 |
| Term | 4.299 (64.21%) | 3385 (65.49%) | 914 (59.89%) |  |
| Postterm | 30 (0.44%) | 19 (0.37%) | 11 (0.72%) |  |
| Birth weight |  |  |  |  |
| Low | 3147 (47.00%) | 2382 (46.08%) | 765 (50.13%) | < 0.001 |
| Adequate | 3424 (51.14%) | 2701 (52.25%) | 723 (47.37%) |  |
| Elevated | 124 (1.85%) | 86 (1.66%) | 38 (2.49%) |  |
| Weight-for-gestational age |  |  |  |  |
| Low | 2247 (33.56%) | 1762 (34.89%) | 485 (31.78%) | 0.244 |
| Adequate | 4228 (63.15%) | 3239 (62.66%) | 989 (64.80%) |  |
| Elevated | 220 (3.28%) | 168 (3.25%) | 52 (3.40%) |  |
| Birth defects |  |  |  |  |
| Yes | 813 (12.14%) | 647 (12.52%) | 166 (10.88%) | 0.093 |
| No | 5882 (87.85%) | 4522 (87.48%) | 1360 (89.12%) |  |
| Neonatal Discharge |  |  |  |  |
| Alive | 6273 (93.69%) | 4859 (94.00%) | 1414 (92.66%) | 0.165 |
| Death | 353 (5.27%) | 259 (5.01%) | 94 (6.16%) |  |
| Transfer | 69 (1.03%) | 51 (0.98%) | 18 (1.18%) |  |

### Factors associated with adolescent pregnancy

Multiple regression analysis showed a significant association between repeated pregnancy and various sociodemographic, obstetric. and perinatal factors. Among the sociodemographic factors, a relationship was identified with the age of 15 to 19 years. Lower educational attainment and the presence of a stable partner. Repeating pregnancy was also associated with pregnancy planning, exposure to violence, the route of cesarean delivery, and preterm birth. No significant association was found with low birth weight (Table 4).

**Table IV.**
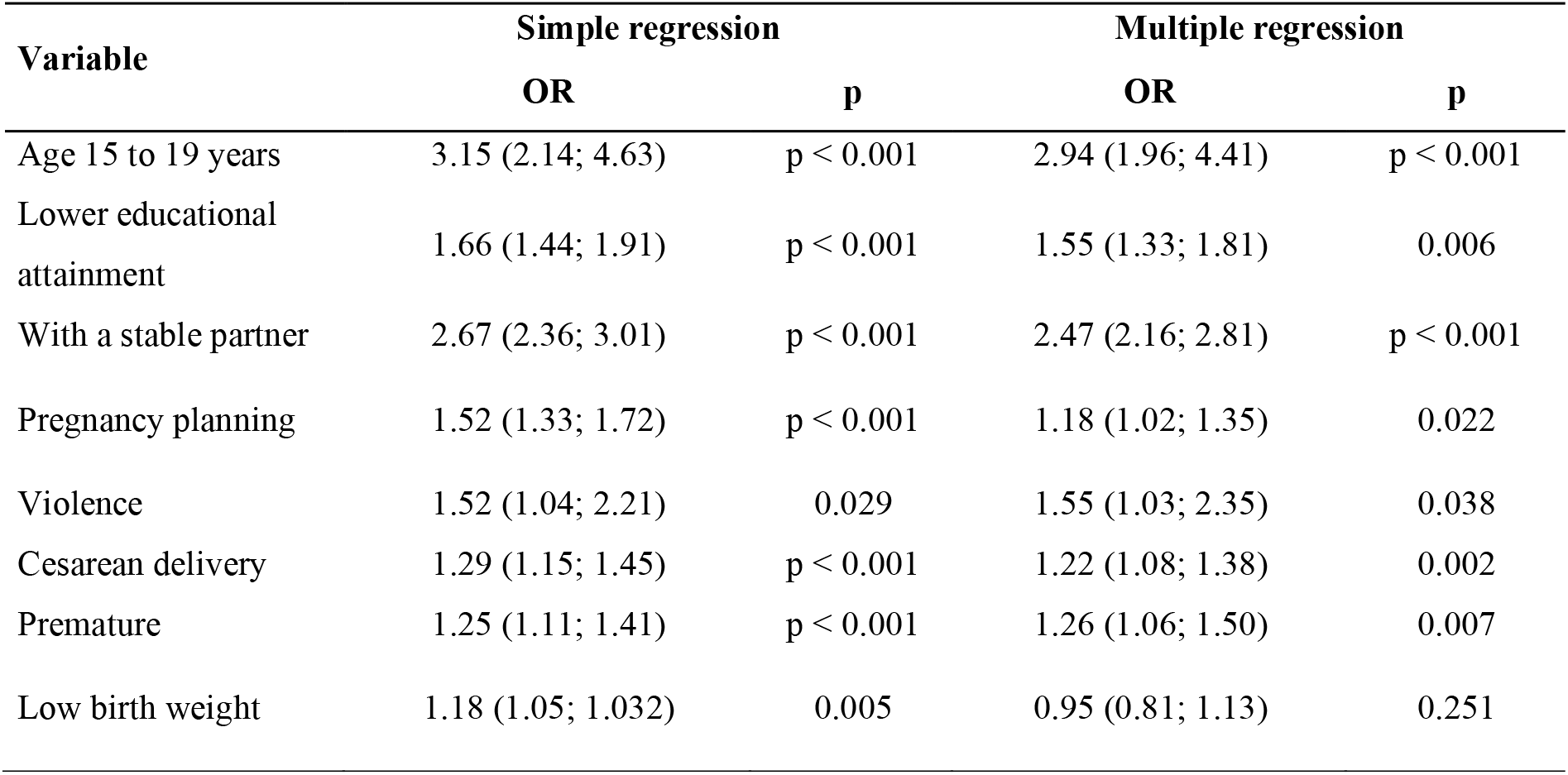
FACTORS ASSOCIATED WITH ADOLESCENT PREGNANCY REPETITION QUITO. ECUADOR. 2009-2022.

## Discussion

The results of this study show that repeated pregnancy in adolescents is a complex phenomenon associated with sociodemographic. pregnancy and perinatal factors, 22.79% of adolescent births were repeat pregnancies, a proportion that is intermediate compared to other studies. In Chile, González (2016) reported 15.6%, while Damle et al. (2015) identified a rate of 35% in the United States.^15,16^

The analysis of the trend of repeated pregnancy in adolescents in Ecuador (2009-2022) showed variations. In 2010. it reached the highest point (>30%). then decreased until 2014. possibly due to the implementation of ENIPLA 2011-2014.^12^ Since 2016, it has stabilized between 15% and 25%, suggesting a stagnation in the downward trend. This pattern is similar in other countries. In Brazil. Monteiro et al. (2023) reported stability with slight reductions (10 to 14 years: 5.0% to 4.7%; 15 to 19 years: 27.8% to 27.3%). In Uganda, Amongin et al. (2020). found that more than half of adolescent girls with a first pregnancy had another birth without rate reduction in the last 30 years.^179^ In Tanzania. Ngoda et al. (2021) showed an increase from 15.8% in 2004-2005 to 18.8% in 2015-2016. reflecting contextual differences.^7^

Stabilization in Ecuador suggests structural barriers that limit the effectiveness of preventive strategies. Despite recent policies such as the National Plan for Sexual and Reproductive Health 2017-2021, the ^18^ Andean Plan for the Prevention of Pregnancy in Adolescents (PLANEA) 2017-2022,^19^ the persistence of the problem indicates the need to strengthen access to effective contraception. sex education. and specialized postpartum care.

In the present study, 98.10% of repeated pregnancies occurred in adolescents aged 15 and 19 years, coinciding with González (2019). Assis et al. (2022) and Ramage et al. (2021). who also identified a higher risk of repeated pregnancy in this age group.^15,20,21^

Low educational attainment was associated with greater repetition of adolescent pregnancy. Diabelková et al. (2023), found that lack of basic or formal education significantly increased the risk (OR = 16.8). Similarly. Zanchi et al. (2017) observed that between four and seven years of schooling doubled the risk. while three years or less tripled it, Monteiro et al. (2023) found that having less than eight years of schooling increased^22,23^ the risk by 64% in adolescents aged 10 to 14 years and by 137% in those aged 15 to 19 years. Maravilla et al. (2017) confirmed the protective effect of schooling. where each additional year of education reduced the risk of repeat pregnancy by 16% (OR = 0.53). ^175^

This study found that having a stable partner is significantly associated with repeat adolescent pregnancy, consistent with previous evidence. Monteiro et al. (2023) reported that being married or in a stable union increased the risk of repeated pregnancy by 96% in adolescents aged 10 to 14 years and by 40% in those aged 15 to 19 years. ^17^ Urquia et al. (2022), found that marriage in children under 18 increased the probability of repeated pregnancy in Ecuador (aOR = 1.99), the United States (aOR = 2.96), and Canada (aOR = 2.17), and with a lower association in Brazil (aOR = 1.09). In Ghana, Boateng et al. (2023) pointed out that cohabitation facilitated the continuity of sexual relations, contributing to the repetition of pregnancy. In addition, many adolescents indicated that the birth of their first child led to premature marriage. perpetuating the risk of new pregnancies.^24, 25^

Although most repeat pregnancies were unplanned, 29.2% were intentionally conceived, suggesting that this phenomenon is not always due to a lack of reproductive control. González (2016) supported this trend by finding that 73.5% of pregnant adolescents did not use contraceptives, Luttges et al. (2021) also highlighted that many young women with repeated pregnancies exhibited a passive attitude towards contraception, which may contribute to repeat pregnancies.^15, 26^

Only 11% of adolescents with repeat pregnancies reported contraceptive use prior to conception, with no significant differences compared to first-time pregnancies. Qasba et al. (2020) highlighted that the use of etonogestrel implant was more effective in preventing repeat pregnancy (3.7%) compared to medroxyprogesterone acetate (22.6%) and short-acting methods (39.1%) (p = 0.01). Not using the implant significantly increased the risk (aOR = 11.8).^27^

Violence in adolescents represents a risk factor for repeated pregnancy. This study evidenced this relationship. supported by Reidy et al (2023). who reported that being a victim of sexual violence increased the number of pregnancies (IRR = 3.32) and children (IRR = 5.43) in a two-year follow-up. The WHO estimates that 24% of adolescent girls aged 15 to 19 have been victims of physical or sexual violence by their partner, and 16% in the last year. Twagirayezu et al. (2024) highlighted that a violent environment contributes to social isolation, school dropout, and economic vulnerability.^28, 29^

Cesarean delivery was significantly associated with repeat pregnancy in adolescents in this study. Similarly, Dimitriu et al. (2019) identified a higher rate of caesarean sections in this group compared to first-time caesarean sections. ^30^

This study confirmed that repeat pregnancy is associated with an increased risk of preterm birth, a finding supported by previous research indicating that adolescent motherhood is associated with this outcome.^31,.32^

This study has limitations inherent to its retrospective and observational design. which prevents causality. However, its methodological strength lies in the large sample and analysis of trends over 14 years (2009-2022).

## Conclusions

Despite the implementation of national public policies, the rate of repeat adolescent pregnancies in Ecuador has remained stable in recent years. This study identified significant associations between repeat pregnancy and sociodemographic factors (maternal age, low educational attainment, and stable partnerships), pregnancy-related variables (planning and exposure to violence), and perinatal outcomes (cesarean delivery and preterm birth). These findings highlight the need to strengthen prevention strategies through comprehensive educational interventions, improved access to reproductive health services, and targeted postpartum follow-up. Additionally, differentiated actions should be prioritized for adolescents at higher risk of pregnancy recurrence.

## Data Availability

All data produced in the present work are contained in the manuscript

## Financing

This study did not receive any funding

